# Comparative effectiveness of ChAdOx1 versus BNT162b2 vaccines against SARS-CoV-2 infections in England and Wales: A cohort analysis using trial emulation in the Virus Watch community data

**DOI:** 10.1101/2021.12.21.21268214

**Authors:** Vincent Grigori Nguyen, Alexei Yavlinsky, Sarah Beale, Susan Hoskins, Vasileios Lampos, Isobel Braithwaite, Thomas E Byrne, Wing Lam Erica Fong, Ellen Fragaszy, Cyril Geismar, Jana Kovar, Annalan M D Navaratnam, Parth Patel, Madhumita Shrotri, Sophie Weber, Andrew C Hayward, Robert W Aldridge, on behalf of the Virus Watch Collaborative

## Abstract

**Introduction:** Infections of SARS-CoV-2 in vaccinated individuals have been increasing globally. Understanding the associations between vaccine type and a post-vaccination infection could help prevent further COVID-19 waves. In this paper, we use trial emulation to understand the impact of a phased introduction of the vaccine in the UK driven by vulnerability and exposure status. We estimate the comparative effectiveness of COVID-19 vaccines (ChAdOx1 versus BNT162b2) against post-vaccination infections of SARS-CoV-2 in a community setting in England and Wales.

**Method:** Trial emulation was conducted by pooling results from six cohorts whose recruitment was staggered between 1st January 2021 and 31st March 2021 and followed until 12th November 2021. Eligibility for each trial was based upon age (18+ at the time of vaccination), without prior signs of infection or an infection within the first 14 days of the first dose. Time from vaccination of ChAdOx1 or BNT162b2 until SARS-CoV-2 infection (positive polymerase chain reaction or lateral flow test after 14 of the vaccination) was modelled using Cox proportional hazards model for each cohort and adjusted for age at vaccination, gender, minority ethnic status, clinically vulnerable status and index of multiple deprivation quintile. For those without SARS-CoV-2 infection during the study period, follow-up was until loss-of-follow-up or end of study (12th November 2021). Pooled hazard ratios were generated using random-effects meta-analysis.

**Results:** Across six cohorts, there were a total of 21,283 participants who were eligible and vaccinated with either ChAdOx1 (n = 13,813) or BNT162b2 (n = 7,470) with a median follow-up time of 266 days (IQR: 235 - 282). By November 12th 2021, 750 (5.4%) adults who had ChAdOx1 as their vaccine experienced a SARS-CoV-2 infection, compared to 296 (4.0%) who had BNT162b2. We found that people who received ChAdOx1 vaccinations had 10.54 per 1000 people higher cumulative incidence for SARS-CoV-2 infection compared to BNT162b2 for infections during a maximum of 315 days of follow-up. When adjusted for age at vaccination, sex, minority ethnic status, index of multiple deprivation, and clinical vulnerability status, we found a pooled adjusted hazard ratio of 1.35 [HR: 1.35, 95%CI: 1.15 - 1.58], demonstrating a 35% increase in SARS-CoV-2 infections in people who received ChAdOx1 compared to BNT162b2.

**Discussion:** We found evidence of greater effectiveness of receiving BNT162b2 compared to ChAdOx1 vaccines against SARS-CoV-2 infection in England and Wales during a time period when Delta became the most prevalent variant of concern. Our findings demonstrate the importance of booster (third) doses to maintain protection and suggest that these should be prioritised to those who received ChAdOx1 as their primary course.

## Introduction

Phase-III randomised controlled trials (RCTs) have demonstrated that the BNT162b2 and ChAdOx1 vaccines are safe and have good efficacy against symptomatic and severe disease (1,2). The UK is one of the first countries to introduce the widespread rollout of the SARs-CoV-2 vaccine compared to other countries. The early introduction of the SARs-CoV-2 vaccine has prevented the deaths and hospitalisations of thousands of individuals (3–5), but England and Wales has recently experienced an uprise in SARs-CoV-2 infections in individuals that have received two vaccines that may be partially attributable to waning vaccine protection and the emergence of the Omicron variant (6).

There are currently no RCTs directly comparing BNT162b2 and ChAdOx1 vaccines to estimate the relative efficacy against COVID-19 infection and disease in the general population. One recently conducted observational study used the target trial design in health and social care workers aged between 18 and 64 and not classed as Clinically Extremely Vulnerable (7). This study found no substantial differences by vaccine type in the incidence of SARS-CoV-2 infection or COVID-19 disease up to 20 weeks after the first vaccination (7). Recent analysis by the UK Health Security Agency (UKHSA) estimates that 20 weeks following the second dose, vaccine effectiveness for ChAdOx1 against infection was 47.3% [95% CI:45.0-49.6] compared to 69.7% [95% CI: 68.7-70.5] for BNT162b2 (8). For hospitalisations due to COVID-19 at 20-weeks, the corresponding vaccine effectiveness was 77.0% [95%CI: 70.3-82.3] for ChadOx1 and 92.7% [95%CI: 90.3-94.6] for BNT162b2. For deaths, vaccine effectiveness at 20 weeks was 78.7 [95% CI 52.7 to 90.4] for ChadOx1 and 90.4 [95% CI 85.1-93.8] for BNT162b2. Overall, these data suggest a faster waning of protection against infection and severe disease for ChAdOx1 compared to BNT162b2 over these longer time periods, but no analyses to date have directly compared BNT162b2 and ChAdOx1 in general population community settings accounting for age and clinical vulnerability and during periods when Delta was the dominant variant of concern.

Due to the United Kingdom’s strategy to prioritise vaccination based upon age, clinical vulnerability and exposure to the virus (for example, frontline healthcare workers) (9), observational studies that do not account for this could experience biases and confounding when comparing individuals who were vaccinated at different points in time. To tackle such biases in this study, methods developed by Hernan and Robins to emulate randomised controlled trials were applied that aim to tackle confounding by indication to appropriately estimate the average treatment effect (10).

In this study, we aim to emulate a randomised controlled trial to estimate the average treatment effect of COVID-19 vaccines (ChAdOx1 or BNT162b2) on SARS-CoV-2 infections in a general population community cohort. Directly estimating the comparative effectiveness of different COVID-19 vaccines aims to reduce the issues of selection bias as a result of access and acceptability to be vaccinated that need to be accounted for in analyses comparing individuals that have been vaccinated to unvaccinated people.

## Method

### Study Design and Setting

The study design used prospective observational data from the Virus Watch Cohort using a target trial emulation study design - a detailed description of the target trial emulation can be found in Table 1. The Virus Watch cohort has been described previously (11). Briefly, households were recruited starting in mid-June 2020 via a number of methods aimed at creating a representative cohort of England and Wales, including postcards or letters sent to the home address, social media and SMS. As of November 2021, 52,677 individuals in 25,309 households had registered to take part. Participating households completed weekly online surveys (reporting a wide range of symptoms, SARS-CoV-2 swab test results and vaccinations) and monthly themed topic surveys. From Autumn 2020, Virus Watch also included a programme of nasopharyngeal swab sample collection and blood collection via venepuncture or finger prick sampling in a subset of 10,000 participants in clinics who are part of the National Institute for Health Research’s Clinical Research Network. From March 2021, blood samples were self-collected by participants using an at-home capillary blood sample collection kit, manufactured by the company Thriva [https://thriva.co/]. Completed kits were returned by participants using pre-paid envelopes and priority postage boxes to UKAS-accredited laboratories for serological testing using Roche’s Elecsys Anti-SARS-CoV-2 assays targeting total immunoglobulin (Ig) to the Nucleocapsid (N) protein or to the receptor-binding domain in the S1 subunit of the Spike protein (S) (Roche Diagnostics, Basel, Switzerland).

**Table 1:** Details of the trial emulation framework

|  | **Ideal Randomised Controlled Trial** | **Trial emulation** |
| --- | --- | --- |
| **Eligibility Criteria** | At least 18 years old<br>No prior SARS-CoV-2 infection<br>No prior SARS-CoV-2 vaccination<br>No SARS-CoV-2 infection between day 0 and day 14 of vaccination to account for immunity development | At least 18 years old when vaccinated<br>No prior SARS-CoV-2 vaccination<br>No reported SARS-CoV-2 infection prior to vaccination date<br>Defined using: PCR, LFT, nucleocapsid antibodies and spike protein before 2021<br>No SARS-CoV-2 infection between day 0 and day 14 of vaccination to account for immunity development |
| **Recruitment period** | 1st January 2021 to 31st March 2021 | 1st January 2021 to 31st March 2021 split by 14-day intervals:<br>01st Jan 2021 - 15th Jan 2021<br>16th Jan 2021 - 30th Jan 2021<br>31st Jan 2021 - 14th Feb 2021<br>15th Feb 2021 - 01st Mar 2021<br>02nd Mar 2021 - 16th Mar 2021<br>17th Mar 2021 - 31st Mar 2021 |
| **Follow-up duration** | From vaccination until 12th November 2021 | From vaccination until 12th November 2021 |
| **Outcome** | 1) Positive PCR test for SARS-CoV-2<br>2) Positive LFT for SARS-CoV-2 | 1) Positive PCR test for SARS-CoV-2 (self-reported or linked)<br>2) Positive LFT for SARS-CoV-2 (self-reported or linked) |
| **Treatments to be compared** | The first dose of ChAdOx1 versus the first dose of BNT162b2 | The first dose of ChAdOx1 versus the first dose of BNT162b2 |
| **Estimand** | Intention to treat | Intention to treat |
| **Analysis plan** | Survival analysis (Kaplan Meier estimator) | Survival analysis (pooled multivariable Cox Proportional hazard models) |

### Participants

Participants from the Virus Watch cohort were considered eligible for the current analyses if they reported having a COVID-19 vaccination (either ChAdOx1 or BNT162b2) between 1st January 2021 to 31st March 2021. As the UK vaccination programme only started vaccinating children under 18 years of age in the second half of 2021, participants under the age of 18 when vaccinated were excluded from these analyses due to their low numbers. Participants who had evidence of SARS-CoV-2 infection prior to their vaccination were excluded in order to examine vaccine and not natural infection-related immunity. Previous SARS-CoV-2 infection was defined using the following: 1) A positive self-reported PCR test, 2) a positive self-reported LFT test, 3) a positive PCR or LFT test from data linkage 4) presence of the nucleocapsid antibody through venous sampling as part of the Virus Watch laboratory sub-cohort or 5) The presence of the Spike antibody prior to December 2020 as these were likely due to natural infection or participation in a vaccination trial. Participants with SARS-CoV-2 infection up to 14 days after receiving their dose were also excluded as we wanted to estimate protection after antibody development.

### Exposure Variables

Vaccination status was self-reported. In the January 11th 2021 and January 18th 2021 Virus Watch questionnaires, participants were asked about their vaccination status retrospectively. From 25 January 2021 onwards, participants were asked weekly for their vaccination status.

### Outcome Variables

We defined the primary outcome as SARS-CoV-2 infection using: 1) a positive self-reported PCR test, 2) a positive self-reported LFT test, or 3) a positive PCR or LFT test from the linked Second Generation Surveillance System (SGSS) data. As we did not link our data on points 1-3 to symptom data, and our outcome may therefore include asymptomatic cases, we refer to our primary outcome as SARS-CoV-2 infection rather than COVID-19 disease for the purposes of this analysis.

### Covariates

Self-reported demographic data included date of birth, sex, and ethnicity. We also included clinically vulnerable status; this was derived from self-reported data on current immunosuppressive therapy, cancer diagnosis (previous or current), and current chronic diseases. Age was calculated based upon the difference between the vaccination date and the reported date of birth. Index of multiple deprivation quintiles was derived based upon Lower Layer Super Output Areas postcodes submitted during registration. Due to small sample sizes (particularly by staggering cohorts), we could not evaluate the impact of geographical region or ethnicity in detail; therefore, we classified ethnicity as “White British” or “Ethnic Minority”.

### Data Sources and Linkage

The primary source of data was the Virus Watch dataset linked to the Second Generation Surveillance System (SGSS), which contains SARS-CoV-2 test results using data from hospitalisations (Pillar 1) and community testing (Pillar 2). Linkage was conducted by NHS Digital with the linkage variables being sent in March 2021. The linkage period for SGSS Pillar 1 encompassed data from March 2020 until August 2021 and from June 2020 until November 2021 for Pillar 2. See Appendix 1 for the linkage period.

### Bias

To estimate the risk of SARs-CoV-2 infection after receiving a COVID-19 vaccine, time-to-event analyses could be conducted. However, evaluating time to SARs-CoV-2 infection may be confounded by the United Kingdom’s strategy to prioritise vaccinations based upon age, clinical vulnerability and exposure to the virus (for example, frontline healthcare workers). To tackle such biases, we used methods developed by Hernan and Robins aim to tackle confounding by indication to appropriately estimate the average treatment effect (12). This approach includes three primary components 1) excluding prevalent users of an intervention to estimate the impact of treatment initiation without the lingering effect of previous treatment, 2) use of an intention to treat analysis as this is the common estimand in randomised controlled trials, and 3) the use of multiple staggered cohorts to appropriately account for “time zero” (or the start of follow-up).

To apply Hernan et al.’s recommendations, this study 1) use eligibility criteria that exclude those who are likely to have protection from SARs-COV-2 (e.g., through prior natural infection), 2) follow individuals from the first vaccination dose and disregard whether further doses were taken (as per recommended to be “fully vaccinated”) or not and 3) stagger the cohort based upon vaccination date. Staggering a single cohort into multiple cohorts aims to produce cohorts that are homogenous in terms of the eligibility criteria that had allowed them to be vaccinated at that point in time. Using staggered cohorts also allows similar individuals to have similar follow-up periods and experience the same COVID-19 public health policies and SARs-CoV-2 reproduction rates at the time of vaccination and throughout their follow-up period. This approach aims to control for the UK’s vaccination prioritisation list but could also mitigate the effects of unmeasured time-varying confounders at the community level, for example, the introduction of new SARS-CoV-2 variants. Therefore, it appropriately accounts for “time zero” (start of follow-up) as it avoids comparisons between individuals who experienced different public health policies and SARs-CoV-2 reproduction rates through time.

### Statistical Analysis

Pooled Cox proportional hazard models were used to estimate the time from vaccination until the primary outcome of SARS-CoV-2 infection, lost to follow-up (latest week of reporting to Virus Watch), or end of study (12th November 2021), whichever was earliest. Cohorts were split based upon the date of their self-reported vaccination with the cohort dates defined in table 2.

**Table 2:**
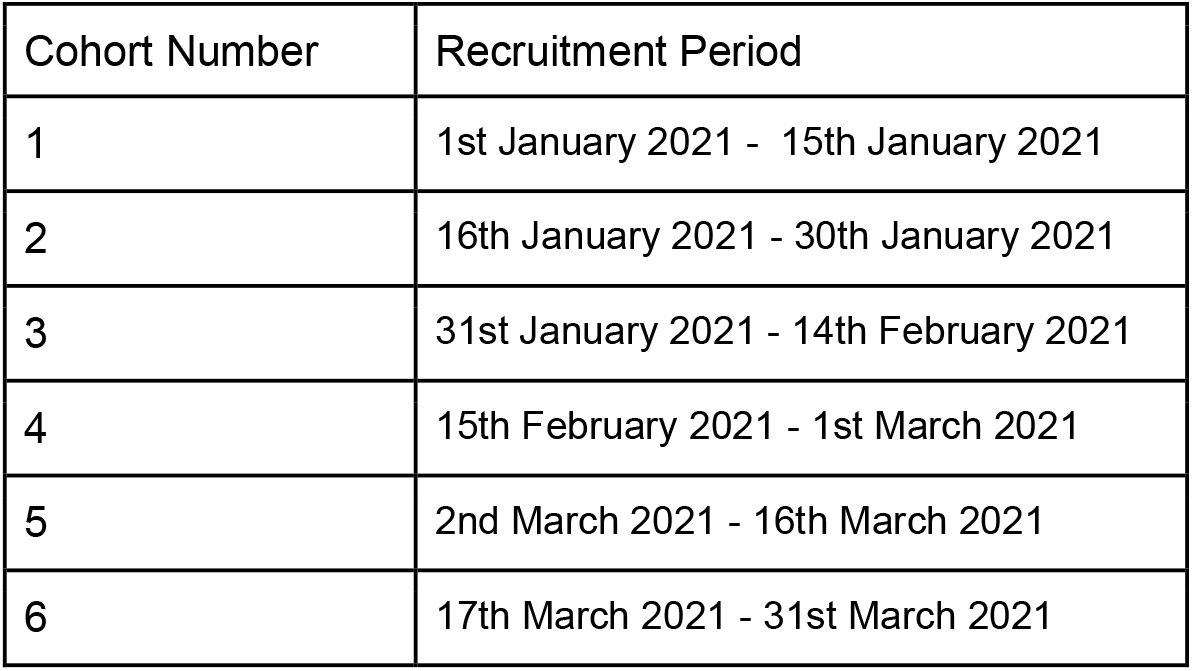
Trial emulation cohorts

| Cohort Number | Recruitment Period |
| --- | --- |
| 1 | 1st January 2021 - 15th January 2021 |
| 2 | 16th January 2021 - 30th January 2021 |
| 3 | 31st January 2021 - 14th February 2021 |
| 4 | 15th February 2021 - 1st March 2021 |
| 5 | 2nd March 2021 - 16th March 2021 |
| 6 | 17th March 2021 - 31st March 2021 |

Multivariable adjustment was conducted using the following variables: age at vaccination, sex, ethnic minority status, index of multiple deprivation quintiles and clinical vulnerable status (clinically vulnerable, clinically extremely vulnerable, or none identified). The models from the six cohorts were then pooled using a random-effects meta-analysis. Full case analysis was conducted for all analyses; therefore, numbers were higher for univariate analysis when compared to multivariable adjustment.

Statistical analysis was conducted using R version 4.0.3,

### Ethical approval

This study has been approved by the Hampstead NHS Health Research Authority Ethics Committee. Ethics approval number - 20/HRA/2320.

## Results

Across the six cohorts, among those who met the eligibility criteria (at least 18 years old at the time of vaccination without prior evidence of a SARs-CoV-2 infection), a total of 21,283 participants were vaccinated between 1st January 2021 to 31st March 2021. The largest recruitment period was between 31st January 2021 to 14th February 2021, with 6,346 participants (3,933 ChAdOx1 versus 2,413 for BNT162b2). The smallest recruitment period was between 1st January 2021 to 15th January 2021, with 1,416 participants (198 ChAdOx1 versus 1,218 for BNT162b2). See figure 1 for the recruitment timeline of the emulated trials.

**Figure 1:**
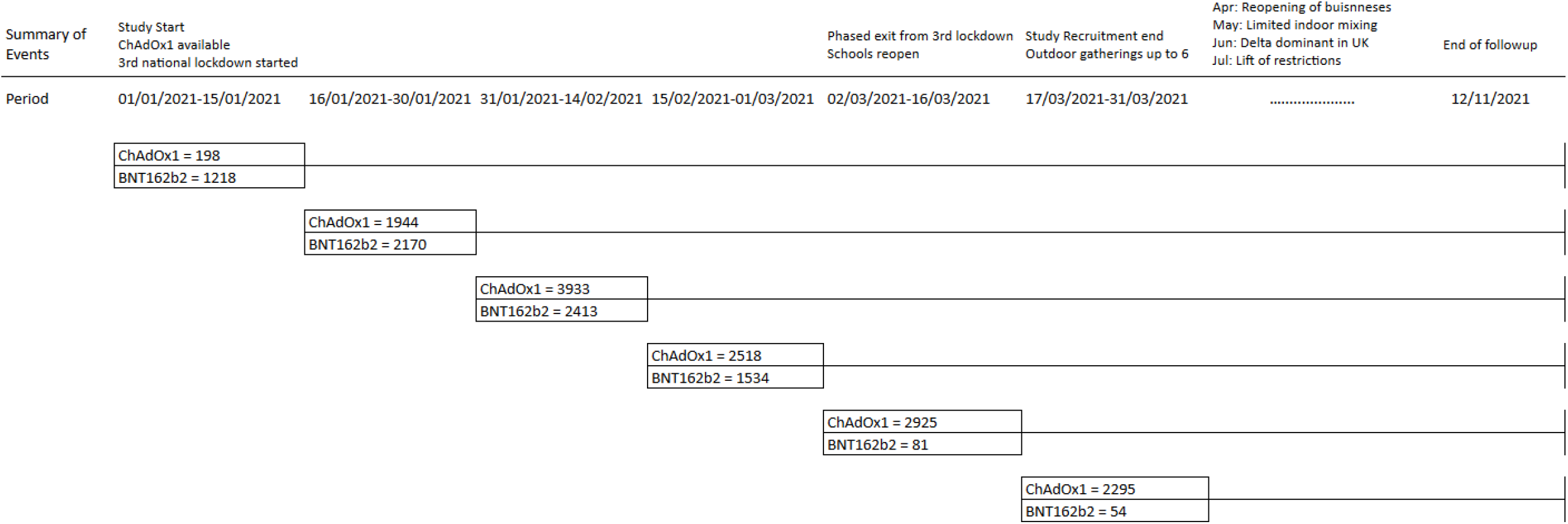
Cohort recruitment numbers and relevant events through time

Across all six cohorts, 13,813 individuals had received the ChAdOx1 vaccine, whilst 7,470 received the BNT162b2 vaccine. Demographic characteristics were broadly similar between ChAdOx1 and BNT162b2, except for clinical vulnerability status and age at vaccination, where BNT162b2 had slightly more clinically vulnerable patients with an older age group (see table 3). Due to the staggering cohort design, it is more appropriate to evaluate the demographics table for each individual trial (see tables 4 to 9). In brief, the median age for both vaccines shows a general decline from trial two onwards; BNT162b2 starts at a median age of 72 (IQR: 63 - 75) at trial two and ends with a median age of 48 (IQR: 42-54) for trial six whilst ChAdOx1 starts with a median age of 73 (IQR: 69-76) at trial two and ends with a median age of 52 (IQR: 48 - 56) by trial six. In terms of clinical vulnerability, for both vaccines, the proportion of those identified as “clinically extremely vulnerable” is the same or lower for each successive trial starting from trial two when compared to the previous trial.

**Table 3:** Baseline demographics for the whole cohort

| Characteristic | N | Pfizer BioNTech,<br>N = 7,470<sup>1</sup> | Oxford AstraZeneca,<br>N = 13,813<sup>1</sup> | p-value<sup>2</sup> |
| --- | --- | --- | --- | --- |
| **Household Region** | 20,598 |  |  | <0.001 |
| East Midlands |  | 666 (9.2%) | 1,232 (9.2%) |  |
| East of England |  | 1,647 (23%) | 3,090 (23%) |  |
| London |  | 919 (13%) | 1,410 (11%) |  |
| North East |  | 303 (4.2%) | 707 (5.3%) |  |
| North West |  | 815 (11%) | 1,434 (11%) |  |
| South East |  | 1,439 (20%) | 2,620 (20%) |  |
| South West |  | 567 (7.9%) | 1,075 (8.0%) |  |
| Wales |  | 160 (2.2%) | 308 (2.3%) |  |
| West Midlands |  | 364 (5.0%) | 797 (6.0%) |  |
| Yorkshire and The Humber |  | 339 (4.7%) | 706 (5.3%) |  |
| Unknown |  | 251 | 434 |  |
| **IMD Quintile** | 20,598 |  |  | 0.5 |
| (Least deprived) 1 |  | 537 (7.4%) | 1,045 (7.8%) |  |
| 2 |  | 1,011 (14%) | 1,930 (14%) |  |
| 3 |  | 1,536 (21%) | 2,774 (21%) |  |
| 4 |  | 1,886 (26%) | 3,557 (27%) |  |
| (Most deprived) 5 |  | 2,249 (31%) | 4,073 (30%) |  |
| Unknown |  | 251 | 434 |  |
| **Sex** | 20,517 |  |  | 0.2 |
| Female |  | 4,078 (57%) | 7,436 (56%) |  |
| Male |  | 3,118 (43%) | 5,885 (44%) |  |
| Unknown |  | 274 | 492 |  |
| **Minority Ethnic Status** | 20,427 |  |  | 0.5 |
| Minority ethnic |  | 704 (9.8%) | 1,259 (9.5%) |  |
| White British |  | 6,469 (90%) | 11,995 (91%) |  |
| Unknown |  | 297 | 559 |  |
| **Clinical vulnerable status** | 21,283 |  |  | <0.001 |
| None identified |  | 4,205 (56%) | 8,612 (62%) |  |
| Clinically extremely vulnerable |  | 1,006 (13%) | 1,507 (11%) |  |
| Clinically vulnerable |  | 2,259 (30%) | 3,694 (27%) |  |
| **Age at vaccination** | 21,283 | 67 (59, 73) | 63 (55, 69) | <0.001 |
| **Post vaccine infection** | 21,283 | 296 (4.0%) | 750 (5.4%) | <0.001 |
| **Follow-up duration** | 21,283 | 276 (259, 291) | 255 (231, 277) | <0.001 |
| <sup>1</sup> n (%); Median (IQR) |  |  |  |  |
| <sup>2</sup> Pearson's Chi-squared test; Wilcoxon rank sum test |  |  |  |  |

**Table 4:** Cohort 1- 1st January 2021 to 15th January 2021

| Characteristic | N | Pfizer BioNTech,<br>N = 1,218<sup>1</sup> | Oxford AstraZeneca,<br>N = 198<sup>1</sup> | p-value<sup>2</sup> |
| --- | --- | --- | --- | --- |
| **Sex** | 1,359 |  |  | 0.3 |
| Female |  | 697 (59%) | 119 (64%) |  |
| Male |  | 475 (41%) | 68 (36%) |  |
| Unknown |  | 46 | 11 |  |
| **Age at vaccination** | 1,416 | 70 (55, 79) | 71 (57, 77) | 0.7 |
| **Minority Ethnic Status** | 1,350 |  |  | 0.6 |
| Minority ethnic |  | 141 (12%) | 25 (13%) |  |
| White British |  | 1,023 (88%) | 161 (87%) |  |
| Unknown |  | 54 | 12 |  |
| **IMD Quintile** | 1,357 |  |  | <0.001 |
| (Least deprived) 1 |  | 64 (5.5%) | 23 (12%) |  |
| 2 |  | 174 (15%) | 18 (9.7%) |  |
| 3 |  | 229 (20%) | 48 (26%) |  |
| 4 |  | 339 (29%) | 43 (23%) |  |
| (Most deprived) 5 |  | 365 (31%) | 54 (29%) |  |
| Unknown |  | 47 | 12 |  |
| **Clinical vulnerable status** | 1,416 |  |  | 0.2 |
| None Identified |  | 743 (61%) | 109 (55%) |  |
| Clinically extremely vulnerable |  | 149 (12%) | 33 (17%) |  |
| Clinically vulnerable |  | 326 (27%) | 56 (28%) |  |
| **Post vaccination infection** | 1,416 | - | - | - |
| **Follow-up duration** | 1,416 | 303 (275, 307) | 302 (241, 304) | <0.001 |
| <sup>1</sup> n (%); Median (IQR) |  |  |  |  |
| <sup>2</sup> Pearson's Chi-squared test; Wilcoxon rank sum test |  |  |  |  |
| -Value suppressed as part of statistical disclosure control |  |  |  |  |

**Table 5:** Cohort 2 - 16th January 2021 - 30 January 2021

| Characteristic | N | Pfizer BioNTech,<br>N = 2,170<sup>1</sup> | Oxford AstraZeneca,<br>N = 1,944<sup>1</sup> | p-value<sup>2</sup> |
| --- | --- | --- | --- | --- |
| **Sex** | 3,963 |  |  | 0.082 |
| Female |  | 1,187 (57%) | 1,008 (54%) |  |
| Male |  | 907 (43%) | 861 (46%) |  |
| Unknown |  | 76 | 75 |  |
| **Age at vaccination** | 4,114 | 72 (63, 75) | 73 (69, 76) | <0.001 |
| **Minority Ethnic Status** | 3,949 |  |  | 0.11 |
| Minority ethnic |  | 198 (9.5%) | 149 (8.0%) |  |
| White British |  | 1,895 (91%) | 1,707 (92%) |  |
| Unknown |  | 77 | 88 |  |
| **IMD Quintile** | 4,001 |  |  | 0.093 |
| (Least deprived) 1 |  | 158 (7.5%) | 126 (6.7%) |  |
| 2 |  | 292 (14%) | 272 (14%) |  |
| 3 |  | 505 (24%) | 390 (21%) |  |
| 4 |  | 537 (25%) | 520 (28%) |  |
| (Most deprived) 5 |  | 623 (29%) | 578 (31%) |  |
| Unknown |  | 55 | 58 |  |
| **Clinical vulnerable status** | 4,114 |  |  | 0.012 |
| None Identified |  | 1,237 (57%) | 1,022 (53%) |  |
| Clinically extremely vulnerable |  | 343 (16%) | 322 (17%) |  |
| Clinically vulnerable |  | 590 (27%) | 600 (31%) |  |
| **Post vaccination infection** | 4,114 | 78 (3.6%) | 81 (4.2%) | 0.3 |
| **Follow-up duration** | 4,114 | 289 (286, 296) | 289 (286, 293) | <0.001 |
| <sup>1</sup> n (%); Median (IQR) |  |  |  |  |
| <sup>2</sup> Pearson's Chi-squared test; Wilcoxon rank-sum test |  |  |  |  |

**Table 6:** Cohort 3 - 31st January 2021 to 14th February 2021

| Characteristic | N | Pfizer BioNTech,<br>N = 2,413<sup>1</sup> | Oxford AstraZeneca,<br>N = 3,933<sup>1</sup> | p-value<sup>2</sup> |
| --- | --- | --- | --- | --- |
| **Sex** | 6,139 |  |  | 0.3 |
| Female |  | 1,317 (57%) | 2,103 (55%) |  |
| Male |  | 1,010 (43%) | 1,709 (45%) |  |
| Unknown |  | 86 | 121 |  |
| **Age at vaccination** | 6,346 | 68 (65, 72) | 69 (65, 72) | 0.6 |
| **Minority Ethnic Status** | 6,115 |  |  | 0.014 |
| Minority ethnic |  | 200 (8.6%) | 263 (6.9%) |  |
| White British |  | 2,116 (91%) | 3,536 (93%) |  |
| Unknown |  | 97 | 134 |  |
| **IMD Quintile** | 6,145 |  |  | 0.14 |
| (Least deprived) 1 |  | 176 (7.6%) | 296 (7.8%) |  |
| 2 |  | 270 (12%) | 459 (12%) |  |
| 3 |  | 476 (20%) | 800 (21%) |  |
| 4 |  | 606 (26%) | 1,071 (28%) |  |
| (Most deprived) 5 |  | 799 (34%) | 1,192 (31%) |  |
| Unknown |  | 86 | 115 |  |
| **Clinical vulnerable status** | 6,346 |  |  | 0.003 |
| None Identified |  | 1,357 (56%) | 2,119 (54%) |  |
| Clinically extremely vulnerable |  | 328 (14%) | 661 (17%) |  |
| Clinically vulnerable |  | 728 (30%) | 1,153 (29%) |  |
| **Post vaccination infection** | 6,346 | 84 (3.5%) | 185 (4.7%) | 0.019 |
| **Follow-up duration** | 6,346 | 275 (272, 282) | 276 (271, 280) | <0.001 |
| <sup>1</sup> n (%); Median (IQR) |  |  |  |  |
| <sup>2</sup> Pearson's Chi-squared test; Wilcoxon rank-sum test |  |  |  |  |

**Table 7:** Cohort 4 - 15th February 2021 to 1st March 2021

| Characteristic | N | Pfizer BioNTech,<br>N = 1,534<sup>1</sup> | Oxford AstraZeneca,<br>N = 2,518<sup>1</sup> | p-value<sup>2</sup> |
| --- | --- | --- | --- | --- |
| **Sex** | 3,904 |  |  | 0.013 |
| Female |  | 805 (54%) | 1,420 (59%) |  |
| Male |  | 673 (46%) | 1,006 (41%) |  |
| Unknown |  | 56 | 92 |  |
| **Age at vaccination** | 4,052 | 60 (54, 64) | 63 (58, 66) | <0.001 |
| **Minority Ethnic Status** | 3,892 |  |  | 0.7 |
| Minority ethnic |  | 151 (10%) | 239 (9.9%) |  |
| White British |  | 1,324 (90%) | 2,178 (90%) |  |
| Unknown |  | 59 | 101 |  |
| **IMD Quintile** | 3,916 |  |  | 0.5 |
| (Least deprived) 1 |  | 123 (8.3%) | 195 (8.0%) |  |
| 2 |  | 246 (17%) | 355 (15%) |  |
| 3 |  | 303 (20%) | 514 (21%) |  |
| 4 |  | 379 (26%) | 631 (26%) |  |
| (Most deprived) 5 |  | 430 (29%) | 740 (30%) |  |
| Unknown |  | 53 | 83 |  |
| **Clinical vulnerable status** | 4,052 |  |  | <0.001 |
| None Identified |  | 775 (51%) | 1,552 (62%) |  |
| Clinically extremely vulnerable |  | 178 (12%) | 240 (9.5%) |  |
| Clinically vulnerable |  | 581 (38%) | 726 (29%) |  |
| **Post vaccination infection** | 4,052 | 75 (4.9%) | 156 (6.2%) | 0.082 |
| **Follow-up duration** | 4,052 | 261 (250, 267) | 260 (252, 267) | 0.045 |
| <sup>1</sup> n (%); Median (IQR) |  |  |  |  |
| <sup>2</sup> Pearson's Chi-squared test; Wilcoxon rank-sum test |  |  |  |  |

**Table 8:**
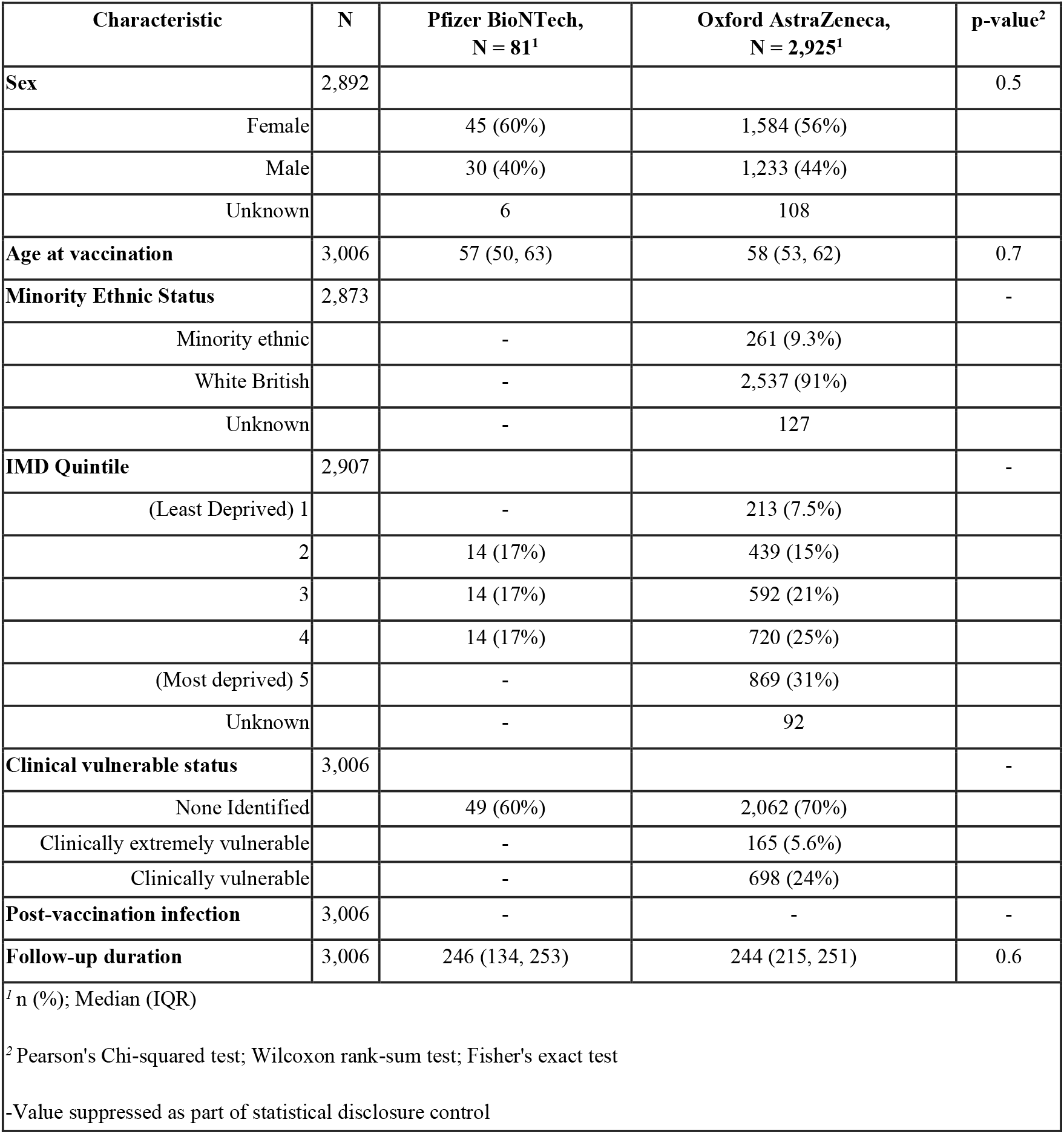
Cohort 5 - 2nd March 2021 to 16th March 2021

| Characteristic | N | Pfizer BioNTech,<br>N = 81<sup>1</sup> | Oxford AstraZeneca,<br>N = 2,925<sup>1</sup> | p-value<sup>2</sup> |
| --- | --- | --- | --- | --- |
| **Sex** | 2,892 |  |  | 0.5 |
| Female |  | 45 (60%) | 1,584 (56%) |  |
| Male |  | 30 (40%) | 1,233 (44%) |  |
| Unknown |  | 6 | 108 |  |
| **Age at vaccination** | 3,006 | 57 (50, 63) | 58 (53, 62) | 0.7 |
| **Minority Ethnic Status** | 2,873 |  |  | - |
| Minority ethnic |  | - | 261 (9.3%) |  |
| White British |  | - | 2,537 (91%) |  |
| Unknown |  | - | 127 |  |
| **IMD Quintile** | 2,907 |  |  | - |
| (Least Deprived) 1 |  | - | 213 (7.5%) |  |
| 2 |  | 14 (17%) | 439 (15%) |  |
| 3 |  | 14 (17%) | 592 (21%) |  |
| 4 |  | 14 (17%) | 720 (25%) |  |
| (Most deprived) 5 |  | - | 869 (31%) |  |
| Unknown |  | - | 92 |  |
| **Clinical vulnerable status** | 3,006 |  |  | - |
| None Identified |  | 49 (60%) | 2,062 (70%) |  |
| Clinically extremely vulnerable |  | - | 165 (5.6%) |  |
| Clinically vulnerable |  | - | 698 (24%) |  |
| **Post-vaccination infection** | 3,006 | - | - | - |
| **Follow-up duration** | 3,006 | 246 (134, 253) | 244 (215, 251) | 0.6 |
| <sup>1</sup> n (%); Median (IQR) |  |  |  |  |
| <sup>2</sup> Pearson's Chi-squared test; Wilcoxon rank-sum test; Fisher's exact test |  |  |  |  |
| -Value suppressed as part of statistical disclosure control |  |  |  |  |

**Table 9:** Cohort 6 - 17th March 2021 to 31st March 2021

| Characteristic | N | Pfizer BioNTech,<br>N = 54<sup>1</sup> | Oxford AstraZeneca,<br>N = 2,295<sup>1</sup> | p-value<sup>2</sup> |
| --- | --- | --- | --- | --- |
| **Sex** | 2,260 |  |  | >0.9 |
| Female |  | 27 (54%) | 1,202 (54%) |  |
| Male |  | 23 (46%) | 1,008 (46%) |  |
| Unknown |  | 4 | 85 |  |
| **Age at vaccination** | 2,349 | 48 (42, 54) | 52 (48, 56) | 0.001 |
| **Minority Ethnic Status** | 2,248 |  |  | 0.6 |
| Minority ethnic |  | - | 322 (15%) |  |
| White British |  | - | 1,876 (85%) |  |
| Unknown |  | - | 97 |  |
| **IMD Quintile** | 2,272 |  |  | - |
| (Least Deprived) 1 |  | - | 192 (8.6%) |  |
| 2 |  | 15 (29%) | 387 (17%) |  |
| 3 |  | 9 (18%) | 430 (19%) |  |
| 4 |  | 11 (22%) | 572 (26%) |  |
| (Most deprived) 5 |  | - | 640 (29%) |  |
| Unknown |  | - | 74 |  |
| **Clinical vulnerable status** | 2,349 |  |  | - |
| None Identified |  | 44 (81%) | 1,748 (76%) |  |
| Clinically extremely vulnerable |  | - | 86 (3.7%) |  |
| Clinically vulnerable |  | - | 461 (20%) |  |
| **Post-vaccination infection** | 2,349 | - | - | - |
| **Follow-up duration** | 2,349 | 230 (198, 233) | 232 (162, 238) | 0.010 |
| <sup>1</sup> n (%); Median (IQR) |  |  |  |  |
| <sup>2</sup> Pearson's Chi-squared test; Wilcoxon rank-sum test; Fisher's exact test |  |  |  |  |
| - Value suppressed as part of statistical disclosure control |  |  |  |  |

Both groups were followed up to a maximum of 315 days (from 1st January 2021 until 12th November 2021), with ChAdOx1 individuals producing a median follow-up duration of 255 days (IQR: 231, 277), whilst BNT162b2 had a median follow-up duration of 276 days (IQR: 259, 291). At 315 days, ChAdOx1 participants experienced a weighted incidence of 51.11 infections per 1,000 vaccinated individuals whilst BNT162b2 participants experienced a weighted incidence of 40.57 infections per 1,000 vaccinated individuals 14-days after being vaccinated; we used the weights for each trial in the random-effects model to calculate these incidences.

### Crude Analysis

We found that ChAdOx1 produces a pooled unadjusted hazard ratio of 1.25 [95% HR: 1.08-1.46; p-value = 0.0038] in the incidence of SARS-CoV-2 infection when compared to BNT162b2 at up to 315 days. For the same time period, the pooled unadjusted hazard ratio for males (reference being female) was 0.90 [95%CI: HR: 0.80 - 1.02; p-value = 0.0996]. The pooled unadjusted hazard ratio per year of age was 0.97 [95%CI: HR: 0.96 - 0.98; p-value = 0.0001].

With the reference variable being “No sign of clinical vulnerability”, the pooled unadjusted hazard ratio for clinically vulnerable was 1.08 [95%: HR: 0.92 - 1.28; p=value = 0.3324] and 1.03 [95%: HR: 0.84 - 1.25; p-value - 0.8014] for “Clinically extremely vulnerable”. See Figure 2 for all unadjusted hazard ratio estimates.

**Figure 2:**
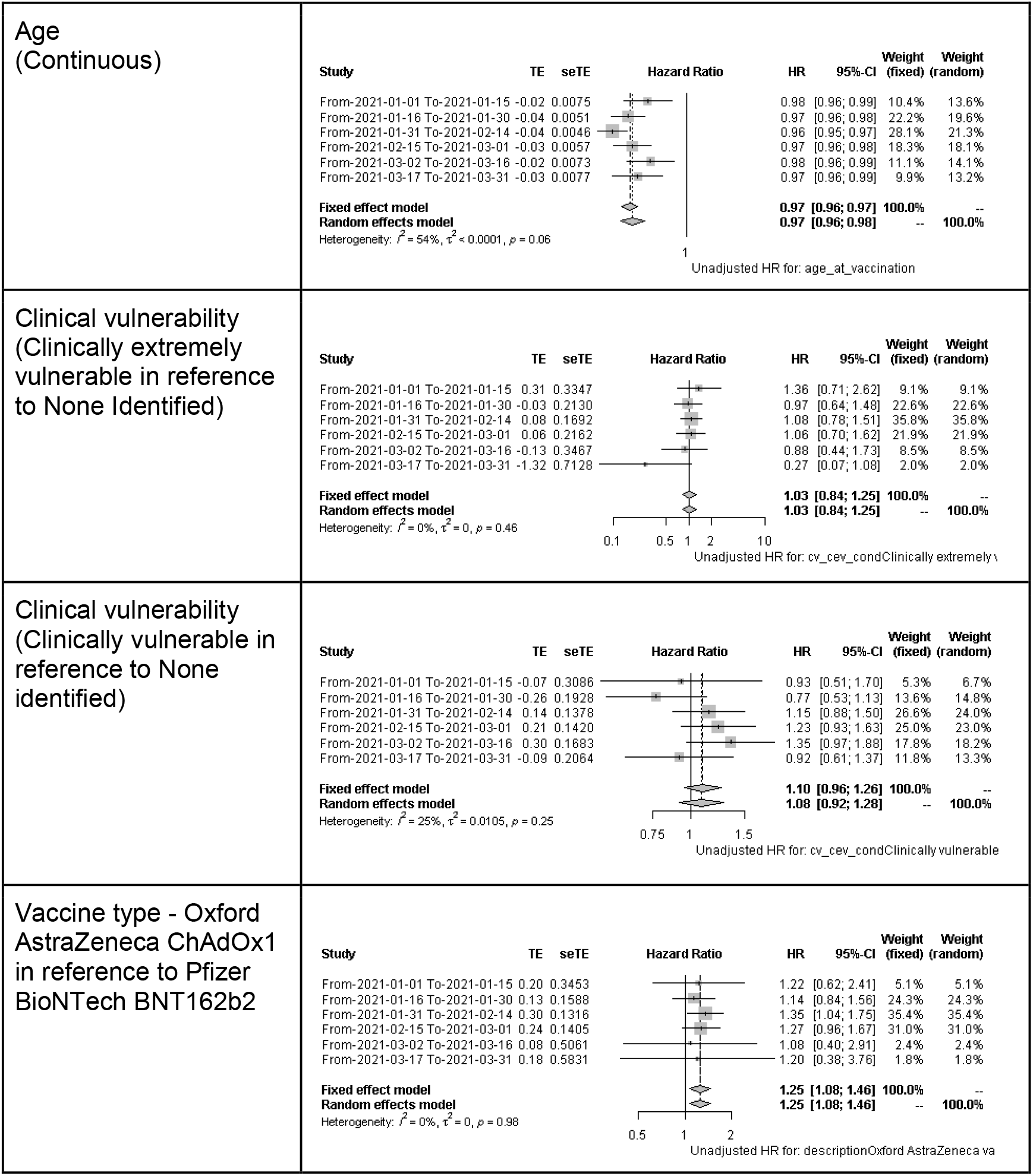
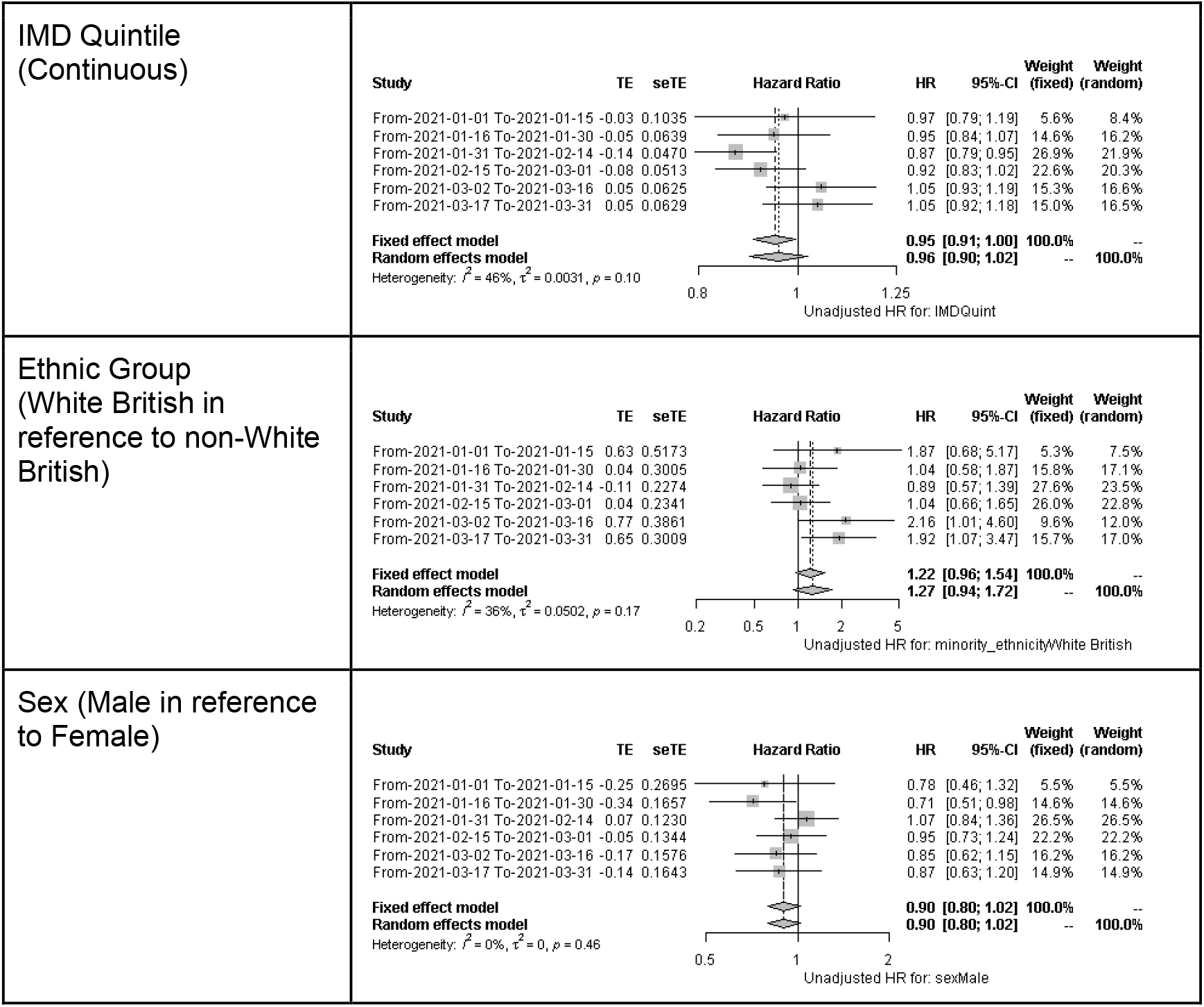
Unadjusted pooled hazard ratios. Hazard Ratio < 1, demonstrates a protective effect. Hazard Ratio = 1 demonstrates no effect. Hazard Ratio > 1, demonstrates an aggravating effect.

### Adjusted Analysis

After adjusting for age at vaccination, clinical vulnerability, IMD quintile, minority ethnic status and sex, the pooled adjusted hazard ratio for ChAdOx1 was 1.35 [95%CI: aHR: 1.15 - 1.58; p-value = 0.0002] suggesting a 35% [95%CI: 15% - 58%] increase in SARS-CoV-2 infection 14 days after vaccination compared to BNT162b2 (see Figure 3)

**Figure 3:**
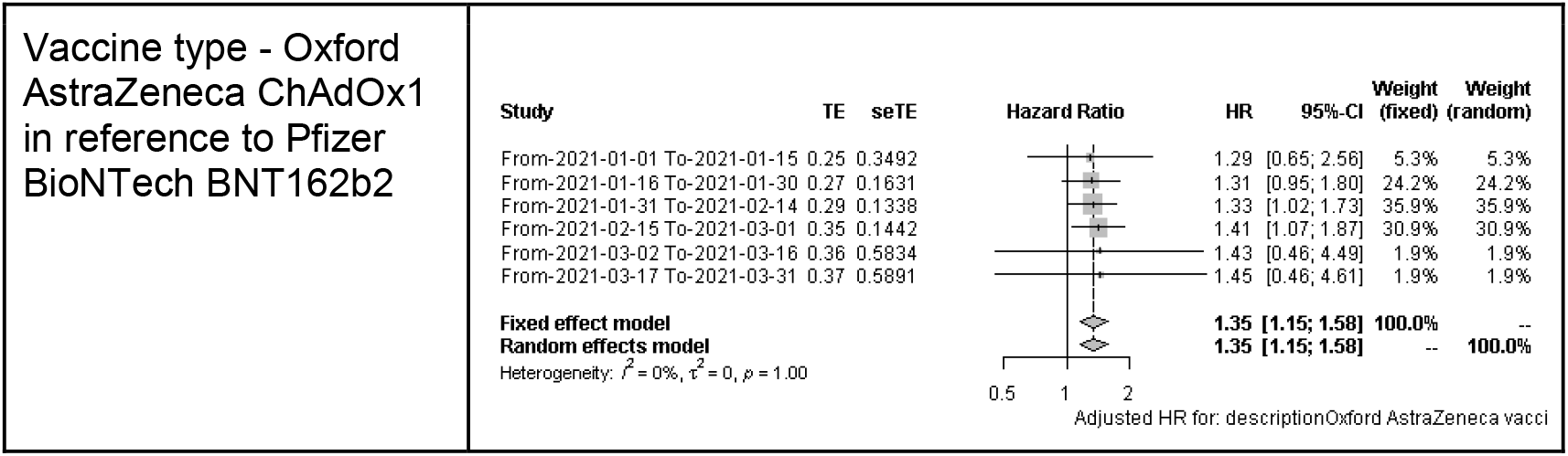
Adjusted Pooled hazard ratio. Hazard Ratio < 1, demonstrates a protective effect. Hazard Ratio = 1 demonstrates no effect. Hazard Ratio > 1, demonstrates an aggravating effect.

## Discussion

Our analysis was conducted in a community cohort of 21,283 people across England and Wales who received their first vaccination between 1st January 2021 to 31st March 2021. We followed people up for risk of SARS-CoV-2 infection between 1st January 2021 and 12th November 2021 and found that people who received ChAdOx1 vaccinations had 10.54 per 1000 people higher cumulative incidence for SARS-CoV-2 infection compared to BNT162b2 for infections during follow-up. We found evidence that people vaccinated with ChAdOx1 had a 35% [95%CI: 15% - 58%; p-value = 0.0002] higher relative risk of SARS-CoV-2 infection compared to BNT162b2 up to 315 days of follow-up after we accounted for differences in demographic and clinical characteristics between our comparison groups as well as the time of vaccination.

Our analysis used a community sample design from across England and Wales, in cohort with diversity in terms of age, sex, and geographical location. We estimated effectiveness in a cohort with a median follow up of eight months after vaccination, and the majority of infections occurred during a period when Delta became the dominant variant in the UK (13). A particular strength of our analysis was our ability to estimate vaccine effectiveness in a cohort that included large numbers of people who were either clinical vulnerable or clinically extremely vulnerable - many previous analyses either did not have such risk factor information or were randomised controlled trials which tended to be conducted in younger and less clinically vulnerable groups. Using this sample, we applied a trial emulation framework to mitigate against confounding by indication. As a result of this study design, we believe our results are more likely to reflect a randomised controlled trial evaluating the same question, and because we are comparing two vaccines, and not unvaccinated individuals, our study design is at less risk of sample bias due to vaccine access and acceptance. The changing median age (which initially increased then decreased after cohort 2) and decreasing clinical vulnerability that we observed in each of our successive cohorts capture the prioritised rollout of the vaccine. We did not include people in nursing homes, which meant that earlier cohorts in the study were more likely to be younger with a clinical risk factor, but after cohort 2, the age-based rollout of the vaccine in the UK becomes prominent. We believe this changing age pattern and decline in extremely clinically vulnerable status across successive cohorts demonstrate that the Virus Watch cohort encompassed the distribution of the vaccine in England and Wales based upon clinical need and vulnerability. We also applied an intention-to-treat analysis as this is the appropriate estimand in randomised controlled trials to account for adherence to treatment, although 88% of our sample had recorded a further SARS-CoV-2 vaccination dose prior to the end of the study.

Our staggered cohort approach has several strengths. First, it enables us to account for the demographic and clinical risk factors of vaccines and make comparisons between similar demographically and clinically similar groups. Second, it can help control for changes in SARS-CoV-2 transmission rates driven by changes in public health policy such as the vaccination efforts (e.g., prioritised distribution and booster campaigns), mask usage, limitations on movement as well the introduction of new SARS-CoV-2 variants into England and Wales(14).

Due to the reliance on self-reported observational studies, there is a risk of inconsistent and inaccurate data recording; however, this was mitigated through linkage to external data sources such as SGSS to complement missing incidence SARS-CoV-2 infections. We measured the risk of SARS-CoV-2 infection as our primary outcome, and whilst this precedes hospitalisation or death, we were not able to look at these more severe outcomes, which is an important limitation of our study. Our use of observational data may mean that there is residual and uncontrolled confounding. Using multiple staggered cohorts reduces the cohort size, and as a result, we had difficulties with analysing certain covariates such as geographical region and ethnicity, which we had to combine into an aggregated category. We did not include occupation or geographical risk in our analyses, and these may result in imbalances in the comparison arms as both risk of exposure to SARS-CoV-2 infection and access to BNT162b2 varied geographically (due to its cold storage requirements) and by occupation (e.g., health and social care workers).

Our findings add to previous analyses which used test negative designs (8), which may be at risk of bias due to eligibility in vaccine rollout programs and when compared to unvaccinated groups, may have important selection biases due to vaccine access and acceptability (15). Our current analysis was conducted using a similar analytical approach to a previous cohort study that emulated a comparative effectiveness trial in health and social care workers up to 20 weeks after the first vaccination (7). This previous study did not find substantial differences in the incidence of SARS-CoV-2 infection or COVID-19 disease up to 20 weeks after vaccination but did suggest a small advantage for BNT162b2 when data were extrapolated beyond the 20 weeks after vaccination.

We found evidence of greater effectiveness of BNT162b2 compared to ChAdOx1 vaccines against SARS-CoV-2 infection in England and Wales, a finding that is consistent with immunological and other assessments of the vaccines (16–18). The waning of immunity for infection and, to a lesser extent, for severe disease, is seen earlier in ChAdOx1 than in BNT162b2 in other UK data and taken together, we believe demonstrate the importance of booster doses to maintain protection and suggest that these should be prioritised to those who received ChAdOx1 as their primary course in the UK and elsewhere.

## Data Availability

All data produced in the present study are not currently available

## Contributors

Study Conceptualisation: VGN, AY, RWA

Formal analysis: VGN, AY, RWA

Project administration: JK, VGN, SB, TB, AMDN, MS, SW.

Data curation: VGN, AMDN, CY, WLEF, MS.

Writing (original draft preparation): VGN, AY, RWA, ACH.

Writing (review and editing): All authors.

Data Access: All authors had full access to the data used in the study.

### Appendix 1

Linkage period: note, during registration, participants were given the opportunity to provide data prior to registration, for example, prior COVID-19 test results

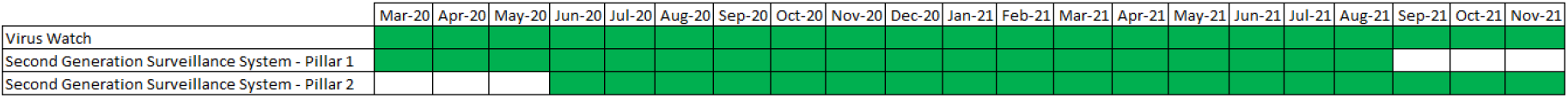

## References

1. Polack FP, Thomas SJ, Kitchin N, Absalon J, Gurtman A, Lockhart S, et al. Safety and Efficacy of the BNT162b2 mRNA Covid-19 Vaccine. N Engl J Med. 2020 Dec 31;383(27):2603–15.

2. Ramasamy MN, Minassian AM, Ewer KJ, Flaxman AL, Folegatti PM, Owens DR, et al. Safety and immunogenicity of ChAdOx1 nCoV-19 vaccine administered in a prime-boost regimen in young and old adults (COV002): a single-blind, randomised, controlled, phase 2/3 trial. The Lancet. 2020 Dec;396(10267):1979–93.

3. UK Health Security Agency. COVID-19 vaccine surveillance report Week 39 [Internet]. 2021 Sep. Available from: https://assets.publishing.service.gov.uk/government/uploads/system/uploads/attachment_data/file/1022238/Vaccine_surveillance_report_-_week_39.pdf

4. Gupta S, Cantor J, Simon KI, Bento AI, Wing C, Whaley CM. Vaccinations Against COVID-19 May Have Averted Up To 140,000 Deaths In The United States: Study examines role of COVID-19 vaccines and deaths averted in the United States. Health Aff (Millwood). 2021 Sep 1;40(9):1465–72.

5. Haas EJ, McLaughlin JM, Khan F, Angulo FJ, Anis E, Lipsitch M, et al. Infections, hospitalisations, and deaths averted via a nationwide vaccination campaign using the Pfizer–BioNTech BNT162b2 mRNA COVID-19 vaccine in Israel: a retrospective surveillance study. Lancet Infect Dis. 2021 Sep;S1473309921005661.

6. UK Health Security Agency. COVID-19 vaccine surveillance report Week 50 [Internet]. 2021 Dec. Available from: https://assets.publishing.service.gov.uk/government/uploads/system/uploads/attachment_data/file/1041593/Vaccine-surveillance-report-week-50.pdf

7. Hulme WJ, Williamson EJ, Green A, Bhaskaran K, McDonald HI, Rentsch CT, et al. Comparative effectiveness of ChAdOx1 versus BNT162b2 COVID-19 vaccines in Health and Social Care workers in England: a cohort study using OpenSAFELY [Internet]. Epidemiology; 2021 Oct [cited 2021 Dec 6]. Available from: http://medrxiv.org/lookup/doi/10.1101/2021.10.13.21264937

8. Andrews N, Tessier E, Stowe J, Gower C, Kirsebom F, Simmons R, et al. Vaccine effectiveness and duration of protection of Comirnaty, Vaxzevria and Spikevax against mild and severe COVID-19 in the UK [Internet]. Epidemiology; 2021 Sep [cited 2021 Dec 6]. Available from: http://medrxiv.org/lookup/doi/10.1101/2021.09.15.21263583

9. JCVI JC on V and I. Priority groups for coronavirus (COVID-19) vaccination: advice from the JCVI, 30 December 2020 [Internet]. 2020 Dec. Available from: https://www.gov.uk/government/publications/priority-groups-for-coronavirus-covid-19-vaccination-advice-from-the-jcvi-30-december-2020

10. Hernán MA, Robins JM. Using Big Data to Emulate a Target Trial When a Randomized Trial Is Not Available: Table 1. Am J Epidemiol. 2016 Apr 15;183(8):758–64.

11. Hayward A, Fragaszy E, Kovar J, Nguyen V, Beale S, Byrne T, et al. Risk factors, symptom reporting, healthcare-seeking behaviour and adherence to public health guidance: protocol for Virus Watch, a prospective community cohort study [Internet]. Infectious Diseases (except HIV/AIDS); 2020 Dec [cited 2021 May 23]. Available from: http://medrxiv.org/lookup/doi/10.1101/2020.12.15.20248254

12. Hernán MA, Alonso A, Logan R, Grodstein F, Michels KB, Willett WC, et al. Observational Studies Analyzed Like Randomized Experiments: An Application to Postmenopausal Hormone Therapy and Coronary Heart Disease. Epidemiology. 2008 Nov;19(6):766–79.

13. UK Health Security Agency. Variants: distribution of case data, 17 December 2021 [Internet]. 2021 Dec. Available from: https://www.gov.uk/government/publications/covid-19-variants-genomically-confirmed-case-numbers/variants-distribution-of-case-data-17-december-2021

14. Singanayagam A, Hakki S, Dunning J, Madon KJ, Crone MA, Koycheva A, et al. Community transmission and viral load kinetics of the SARS-CoV-2 delta (B.1.617.2) variant in vaccinated and unvaccinated individuals in the UK: a prospective, longitudinal, cohort study. Lancet Infect Dis. 2021 Oct;S1473309921006484.

15. Lopez Bernal J, Andrews N, Gower C, Robertson C, Stowe J, Tessier E, et al. Effectiveness of the Pfizer-BioNTech and Oxford-AstraZeneca vaccines on covid-19 related symptoms, hospital admissions, and mortality in older adults in England: test negative case-control study. BMJ. 2021 May 13;n1088.

16. Aldridge RW, Yavlinsky A, Nguyen V, Eyre MT, Shrotri M, Navaratnam AMD, et al. Waning of SARS-CoV-2 antibodies targeting the Spike protein in individuals post second dose of ChAdOx1 and BNT162b2 COVID-19 vaccines and risk of breakthrough infections: analysis of the Virus Watch community cohort [Internet]. Infectious Diseases (except HIV/AIDS); 2021 Nov [cited 2021 Dec 7]. Available from: http://medrxiv.org/lookup/doi/10.1101/2021.11.05.21265968

17. Shrotri M, Navaratnam AMD, Nguyen V, Byrne T, Geismar C, Fragaszy E, et al. Spike-antibody waning after second dose of BNT162b2 or ChAdOx1. The Lancet. 2021 Jul;398(10298):385–7.

18. Katikireddi SV, Cerqueira-Silva T, Vasileiou E, Robertson C, Amele S, Pan J, et al. Two-dose ChAdOx1 nCoV-19 vaccine protection against COVID-19 hospital admissions and deaths over time: a retrospective, population-based cohort study in Scotland and Brazil. The Lancet. 2021 Dec;S0140673621027549.

